# Global effects in fMRI reveal brain markers of state and trait anxiety

**DOI:** 10.1101/2025.07.15.25331571

**Authors:** Kimberly Rogge-Obando, Terra Lee, Caroline G. Martin, Kamalpreet Kaur, Yamin Li, Jeffrey M. Harding, Shiyu Wang, Richard Song, Ruoqi Yang, Rithwik Guntaka, Sarah E. Goodale, Roza G. Bayrak, Lucina Q. Uddin, Martin Walter, Jeremy Hogeveen, Catie Chang

## Abstract

**Background:** To personalize the diagnosis and treatment of anxiety, there is a need to identify biological constructs that underlie self-reported symptoms. Notably, physiological responses and altered levels of arousal are constituents of anxiety and have widespread (“global”) effects on fMRI signals across the brain. Therefore, fMRI signatures of global cortical arousal and autonomic physiological responses may provide valuable neuroimaging biomarkers of anxiety. Additionally, these effects may also contribute to relationships observed between large scale network dynamics and anxiety level.

**Methods:** Drawing upon data from a large community sample of 543 subjects (F= 369, M=174) we examine whether the global mean fMRI signal, and a data-driven estimate of cortical arousal effects in fMRI, relate to state and trait anxiety. Additionally, we investigate if autonomic physiological measures (heart rate) in fMRI patterns relate to state and trait anxiety in a subsample of these subjects (240 subjects; F=154, M=86). Finally, we investigate if these three global fMRI effects influence the relationship between functional brain network connectivity and state and trait anxiety.

**Results:** We observe that the spatial patterns of the global mean fMRI signal and the cortical arousal-related fMRI signal related to both state and trait anxiety. These results support current theories that cortical arousal is closely tied to the anxious experience. Additionally, we observe that global component regression had variable effects on the relationship between anxiety and brain networks.

**Conclusions:** These findings suggest that global effects in fMRI signals hold valuable information about both state and trait anxiety. These observations also underscore the importance of understanding global fMRI effects as a source of information as opposed to a confound.

## Introduction

In 2019, approximately 301 million individuals were diagnosed with an anxiety disorder^1,2^. The current practice for diagnosing an anxiety disorder relies on the Diagnostic of Statistical Manual of Mental Disorders^3^, which is based on an individual’s behavior. Yet, behavioral assessments can be subjective, leading to inconsistent and/or inaccurate diagnoses. To address this limitation, the National Institute of Mental Health initiated the Research Domain Criteria (RDoC) to foster research into fundamental psychological or biological systems to understand mental illnesses more thoroughly.

A widespread approach for investigating biomarkers of anxiety centers on the functional connectivity of large-scale brain networks measured with functional magnetic resonance imaging (fMRI). Overall, large-scale network connectivity tends to show a weak association with anxiety^4^. Nonetheless, multiple studies have reported that the functional connectivity within and between two major brain networks – the salience network^5,6^ and the default-mode network^7^– relates to inter-individual differences in state and trait anxiety^8–10^. However, the precise relationship between brain network connectivity and anxiety appears to be complex. For instance, the inter- and intra-network connectivity of the salience network and the default mode network has been observed in some studies to increase^11,12^ but in others to decrease ^8,11,13^ as anxiety increases. These inconsistencies motivate a continued search for more robust biological markers of anxiety.

In separate lines of work, anxiety has been closely linked with measures of autonomic physiology (e.g., heart rate)^14–16^ and cortical arousal^17^. However, these processes have been under-explored in the context of neuroimaging biomarkers of anxiety. Fluctuations in heart rate ^18–20^ and cortical arousal^21–23^ are known to manifest in fMRI signals, exhibiting widespread (“global”) correlations. It is therefore possible that these effects may help to identify stronger biomarkers of anxiety and reveal brain regions that relate physiological or cortical arousal to anxiety.

While fMRI global components are often considered to be artifactual, growing evidence suggests that they may hold valuable information related to anxiety^20,24–28^. For example, studies measuring heart rate variability during fMRI have identified brain-physiological interactions in several areas that related to anxiety^20,24,25^. The average fMRI signal across the brain (“global mean signal”), which captures both arousal and heart rate, was found to be expressed in brain regions closely tied with gene expression and neurotransmitters crucial to trait anxiety^28^. fMRI studies of cortical arousal have been limited, as validated arousal measures are not commonly acquired during fMRI. However, approaches for estimating fMRI cortical arousal components have been developed^22,29,30^, and can be leveraged to study the association between fMRI arousal and anxiety. Overall, investigating fMRI patterns of heart rate or cortical arousal related to anxiety would take a critical step toward illuminating novel clinical brain markers and treatment targets.

Here, we conducted an exploratory analysis upon a large set of individuals from the Enhanced Nathan Kline Institute-Rockland Sample^31^ to determine if global fMRI components may provide brain-based markers of state or trait anxiety. Although this population was not clinical, our study provides foundational knowledge and methodology for future investigations in clinical populations. We focus on three global fMRI components: (1) fMRI correlates of heart rate, (2) the global mean signal, and (3) a data-driven estimate of cortical arousal-dependent fMRI fluctuations that has been replicated across species and different sites^22,29,30^.

As a second goal, we examine whether regressing global components from fMRI network signals would impact associations between brain network connectivity and anxiety. We focused on three major networks: default mode network, salience network, and central executive network. The central executive network, which interacts with the default-mode and salience networks within a triple-network model^32^, has been found to be abnormal in a range of anxiety disorders^11^, motivating its inclusion. Beyond their potential link with anxiety ^8–10^ interactions between these networks may also be sensitive to arousal^33,34^. We hypothesized that salience network activity would relate to heart rate fluctuations, and that relationships between functional connectivity and both state and trait anxiety would diminish after regressing out global components.

Together, these complementary analyses aim to 1) provide new insight into the value of brain-wide physiological and cortical arousal signatures as markers of anxiety, and 2) inform how interactions between major brain networks and anxiety are impacted by modeling global fMRI effects.

## Methods and Materials

### Data Acquisition

Data were drawn from the Enhanced Nathan Kline Institute-Rockland Sample (NKI-RS;^31^). The institutional review board of the NKI approved data collection, and participants gave informed consent. Resting-state fMRI (EPI sequence, TR = 1400 ms, duration = 10 min, voxel size = 2.0 mm isotropic, FA = 65°, FOV = 224 mm) and a high-resolution anatomic scan (MPRAGE sequence, TR/TE = 1900/2.52 ms, FA = 9°, thickness = 1.0 mm, slices = 192, matrix = 256 × 256, FOV = 250 mm) were acquired. Cardiac (via photoplethysmography; PPG) data were recorded during the resting-state fMRI scan. A total of 543 subjects that had fMRI and State-Trait Anxiety Inventory scores (F = 369, M = 174) were included. All 543 subjects completed the State-Trait Anxiety Inventory (STAI) approximately one day before the MRI scan. Our analyses used the T-scored values as obtained per the guidelines of the STAI scoring manual. Subject demographics are depicted in **Table 1**. From this cohort, 240 subjects (F = 154, M = 86) that had adequate heart rate (HR) quality were used in analyses that required heart rate. Two trained researchers manually inspected the raw PPG data and the derived HR measures (**Supplementary Figure 1**).

**Table 1.** Demographic information and state/trait anxiety scores.

| **Characteristic** | **All subjects (N=543)** | **High Quality HR Subjects (N=240)** |
| --- | --- | --- |
| **Age** | 48.1 ($\pm 17.4$) | 43.8 ($\pm 16.7$) |
| **Sex** | 369/174 (F/M) | 154/86 (F/M) |
| **Ethnicity** | 480/63<br>(Not Hispanic, Latino,<br>Spanish/Hispanic, Latino,<br>Spanish) | 206/34<br>(Not Hispanic, Latino,<br>Spanish/Hispanic, Latino, Spanish) |
| **State (t-scores)** | 46.8 ($\pm 10.2$) | 45.3 ($\pm 9.0$) |
| **Trait (t-scores)** | 50.7 ($\pm 11.6$) | 49.1 ($\pm 10.4$) |
| **Potential STAI Clinical Anxiety Population** | 97/446 (Clinical/Non-clinical) | 32/208 (Clinical/Non-clinical) |

Demographics and State and Trait Anxiety Inventory (STAI) t-scores for the indicated subsets of participants used in this study. Values in the table indicate mean ( ± standard deviation). STAI approximate Clinical Anxiety Ranges were calculated using the raw state score<sup>76,77</sup>. HR: heart rate.

### MRI Data Preprocessing

The T_1_ anatomic images underwent brain extraction (FSL *bet*), reduction of spatial bias field inhomogeneities (AFNI *3dUnifize*), and a nonlinear transformation between the T_1_ image and MNI152 space (ANTS). The fMRI data first underwent motion coregistration (FSL *mcflirt*) followed by ICA FIX^54^ to correct for motion and scanner artifacts. For FIX, we constructed a study-specific training set using 25 subjects from the NKI dataset with a balanced age distribution. The fMRI data were then registered to the associated T_1_ image with an affine transformation (FSL *epi_reg*), followed by alignment to MNI152 space via the previously estimated nonlinear transformation between T_1_ and MNI152 space. Spatial blurring was then performed (AFNI *3dmerge*; FWHM of 3mm), and slow scanner drifts were reduced by removing polynomials up to a 4th order (AFNI *3dDetrend*). We additionally regressed out six rigid-body head motion parameters and their derivatives using MATLAB.

### Heart Rate Preprocessing

To calculate HR, we first band-pass filtered the photoplethysmography data with a second-order Butterworth filter (0.5-2 Hz), detected peaks with a minimum height of 5% of the interquartile range, and calculated the time between successive peaks as the inter-beat interval (IBI). Although scans having high-quality PPG data had been selected for analysis, some of these nonetheless contained transient, fixable artifacts, and we interpolated over the few instances when these occurred. We then derived HR as the inverse of the median IBI per minute within successive 6-s sliding windows centered on the time of each fMRI volume acquisition (i.e., at each TR)^35,36^.

### Deriving Global fMRI Components

For each subject, the fMRI global mean signal was calculated as the average time-series across all voxels in the brain. A voxel-wise spatial map, corresponding to the strength at which the global mean signal is expressed at each voxel, was derived by projecting (via linear regression) the global mean signal onto the time-series of each voxel and extracting the corresponding regression coefficient (beta) at each voxel.

Since the mapping between HR and fMRI cannot be captured by a simple correlation^35,37^, we used basis functions ^56^ to compute five HR regressors after removing polynomial trends up to a 4th order (matching the detrending step of the fMRI data). We computed a map capturing HR effects across the brain by calculating the percentage of temporal variance explained by these HR regressors in each voxel’s fMRI time-series.

Lastly, we used a template-based approach for deriving arousal effects in fMRI^22,29,30^. Briefly, an fMRI Arousal Index (FAI) time-series was derived by first z-scoring each voxel’s fMRI time-series, and then spatially correlating the template map derived in Goodale et al., 2021 onto the fMRI volume at each TR. In addition to this FAI time-series, which provides an estimate of arousal fluctuations across the scan, a voxel-wise spatial map – corresponding to the strength at which this FAI is expressed at each voxel – was derived by projecting (via linear regression) onto the FAI onto the time-series of each voxel and extracting the corresponding regression coefficient (beta) at each voxel.

### Relating Global Components to State and Trait Anxiety Measures

To identify brain regions in which the expression of each global component is associated with anxiety measures, we used a GLM (implemented in FSL *randomise*) to relate subject-specific global signal maps, heart rate variation maps, and FAI maps to state and trait anxiety scores. Correction for multiple comparisons was carried out using Threshold Free Cluster Enhancement with 5000 permutations. We also related temporal features of each global component to anxiety measures, including the standard deviation of the FAI time-series (which serves as an estimated drowsiness value^22^) and of the global mean signal. Age, sex, and ethnicity were included as covariates in these analyses, since they can significantly impact fMRI signals, including the global mean signal and heart rate^38–41^. Given the wide age range of the sample, we additionally recomputed the analyses without correcting for age as well as after excluding subjects older than 55 years^39^.

### Deriving Resting-State Networks and Computing Functional Connectivity

Group-level independent component analysis (ICA), implemented in FSL Melodic, was applied to derive 40 components from a randomly selected group of 50 subjects. These components were visually inspected to identify five networks of interest: dorsal default mode network (DDMN), ventral default mode network (VDMN), salience network (SAL), left central executive network (LCEN), and right central executive network (RCEN). Next, dual regression^58,59^ was used with all 40 components to derive subject-specific time-series and spatial maps for each component. Pairwise functional connectivity was then computed amongst the five networks of interest, using Pearson correlation.

### Relating Global Component Signals to Resting-State Network Signals

To understand the relationship between the time-series of the global components and those of the functional networks, we correlated the FAI and global mean signals to each network of interest. For the subset of 240 subjects with sufficient heart rate signal quality, we also calculated lagged cross-correlations between heart rate and each network of interest.

### Relating Functional Connectivity to State and Trait Anxiety Measures

We constructed a mixed model that relates functional connectivity strength with state or trait anxiety score, covarying for age, sex, and ethnicity. We repeated this analysis after regressing out one or more global components from the network time-series. We replicated these procedures for the subset of 240 subjects with high-quality heart rate data.

## Results

### Interactions between global fMRI components and state and trait anxiety

The spatial maps of the FAI and of the global mean signal exhibited significant relationships (p<0.05, corrected) to state and trait anxiety **Figure 1(B,C,E,F)**. The global mean signal has an inverse relationship with the FAI (^22,29,42^; see also next section), which accounts for the differences in sign between their respective statistical maps. No significant clusters were found for the heart-rate spatial maps. When conducting this analysis with a subset of individuals between ages 18-55, only associations between state anxiety and FAI spatial maps showed significant clusters (**Supplementary Figure 2**).

**Figure 1.**
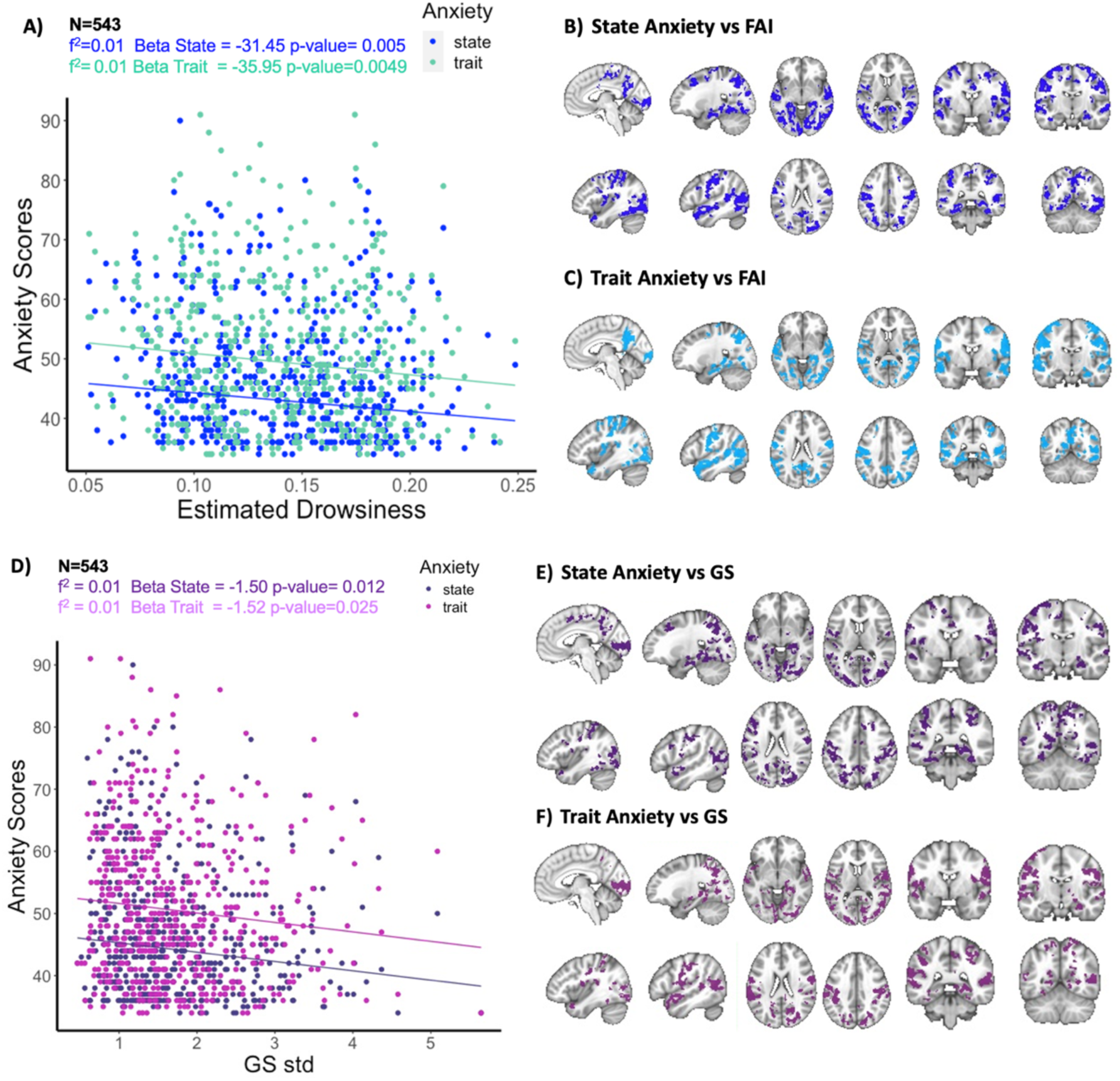
Relating spatial and dynamic aspects of global brain arousal effects to state and trait anxiety. (A) Relationship between anxiety scores and the estimated drowsiness level during the fMRI scans, covarying for age, sex, and ethnicity. (B, C) Brain regions where subject-specific spatial maps associated with FAI were significantly related to state or trait anxiety, covarying for age, sex, and ethnicity. Clusters indicate regions where there is a weaker association with FAI as anxiety increases; no clusters were found to exhibit the opposite relationship. D) Relationship between the standard deviation of the global mean signal and anxiety scores while correcting for age, sex, and ethnicity. (E, F) Brain regions where subject-specific spatial maps associated with the global mean signal were significantly related to state or trait anxiety, covarying for age, sex, and ethnicity. Clusters indicate regions where there is less association with the indicated global signal as anxiety increases. All analyses in this figure are based on n=543 subjects, and the significance of clusters in the spatial maps was determined at a threshold of p<0.05, corrected for multiple comparisons using threshold-free cluster enhancement.

The estimated drowsiness level (standard deviation of the FAI time-series) exhibited a significant relationship with both state and trait anxiety (β= −31.5, p= 0.005; β= −35.9, p= 0.005; **Figure 1(A,D)**. The standard deviation of the global mean signal was also related to state and trait anxiety, albeit with a weaker effect size (β= −1.5, p= 0.012; β= −1.52, p= 0.025). We then examined effects of age, finding that both estimated drowsiness (r=-0.25, p<0.001) and the standard deviation of the global signal (r=-0.39, p=<0.001) significantly correlated with age. However, age did not relate to either state (r=0.06, p=0.14) or trait (r=0.02, p=0.61) anxiety measures, suggesting it does not drive the relationships between global fMRI effects and anxiety. Nonetheless, as a secondary check, we replicated the analysis after removing subjects older than 55. We found that the strength of associations diminished, although the direction of effects was preserved (**Supplementary Figure 2**).

### Relationship between global component dynamics and network dynamics

Correlations between the time-series of global components and functional networks are depicted in **Figure 2(B,C)**, and correlations between global components are shown in **Supplementary Figure 3**. Given the inverse relationship between the FAI and the global mean signal, correlations with the FAI component were multiplied by −1 for display. For the set of 543 subjects, we observed that the 5 networks of interest generally exhibited a stronger median correlation with the global mean signal compared to the FAI signal. For the subset of 240 subjects with heart-rate measures, the global mean signal or heart rate tended to exhibit the strongest median correlations with network signals.

**Figure 2.**
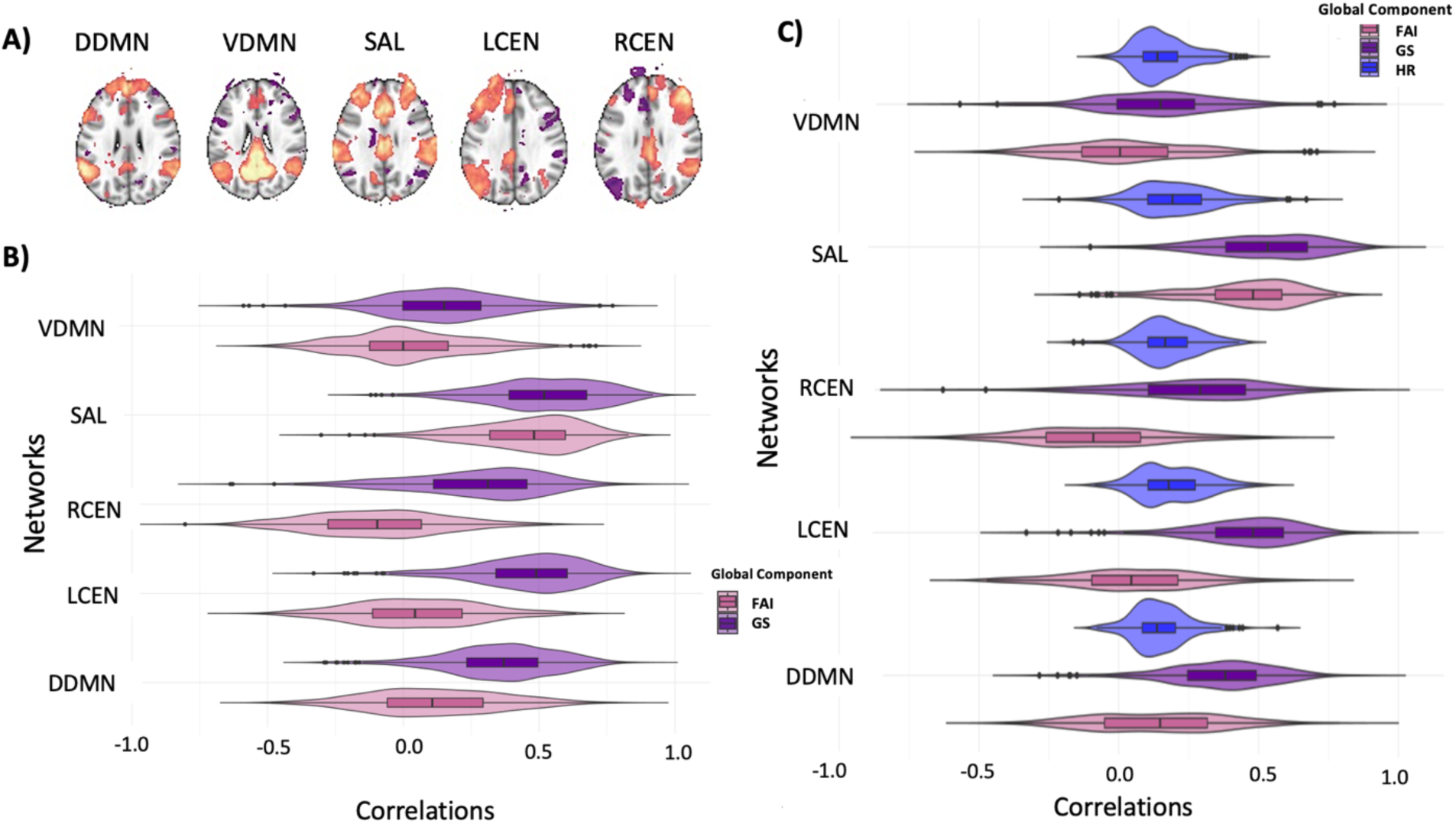
Relating large-scale networks to global components. (A) ICA components identified as the five networks of interest: dorsal default mode network (DDMN), ventral default mode network (VDMN), salience network (SAL), left central executive network (LCEN), and right central executive network (RCEN). Distribution of correlations between the indicated global components and each network of interest, shown for (B) 543 subjects, and (C) the set of 240 subjects with clean heart-rate measures. The global components used in the analysis of 543 subjects included only FAI and the global mean signal, while those of the 240-subject subset additionally included heart rate. Since FAI and GS exhibited negative correlations with one another, the sign of the FAI signal was reversed here for display.

### Impact of regressing global components on the association between functional connectivity and state and trait anxiety

Relationships between network connectivity and anxiety measures, with and without regressing global components out of the network signals, are depicted in **Table 3**. We identified a set of pairwise network connections whose relationship with either state or trait anxiety exceeded p<0.05 (uncorrected). Before regressing out any global components, the connectivity between VDMN-SAL and RCEN-SAL survived this threshold for both state and trait anxiety. When FAI time-series were regressed out, the relationships between RCEN-SAL and state anxiety, and VDMN-SAL and trait anxiety, no longer survived this threshold.

**Table 2.** Impact of regressing global fMRI components on the association between functional connectivity and anxiety.

| Number of Subjects | N=543 |  |  |  |  |  |  |  | N=240 |  |
| --- | --- | --- | --- | --- | --- | --- | --- | --- | --- | --- |
| Psychometric | State Anxiety |  |  |  | Trait Anxiety |  |  |  | Trait Anxiety |  |
| Network Connectivity | VDMN-SAL |  | RCEN-SAL |  | VDMN-SAL |  | RCEN-SAL |  | DDMN-RCEN |  |
| Non-Regressed | * | (-) | * | (-) | * | (-) | * | (-) | * | (+) |
|  | 0.023 |  | 0.039 |  | 0.041 |  | 0.012 |  | 0.033 |  |
| HR Regressed |  |  |  |  |  |  |  |  | * | (+) |
|  |  |  |  |  |  |  |  |  | 0.034 |  |
| FAI Regressed | * | (-) | 0.051 | (-) | 0.10 | (-) | ** | (-) | * | (+) |
|  | 0.017 |  |  |  |  |  | 0.010 |  | 0.034 |  |
| GS Regressed | * | (-) | * | (-) | 0.24 | (-) | * | (-) | 0.064 | (+) |
|  | 0.035 |  | 0.035 |  |  |  | 0.012 |  |  |  |

Association between pairwise functional connectivity and psychometric variables (state or trait anxiety). Entries in the table represent uncorrected p-values before and after the indicated global components were regressed out of the network time courses, together with the sign of their respective beta value. Global components consisted of the global mean signal (GS), an fMRI-derived arousal index (FAI), and heart rate (HR). \*\*p<0.01 uncorrected, \*p<0.05 uncorrected.

**Table 3.**
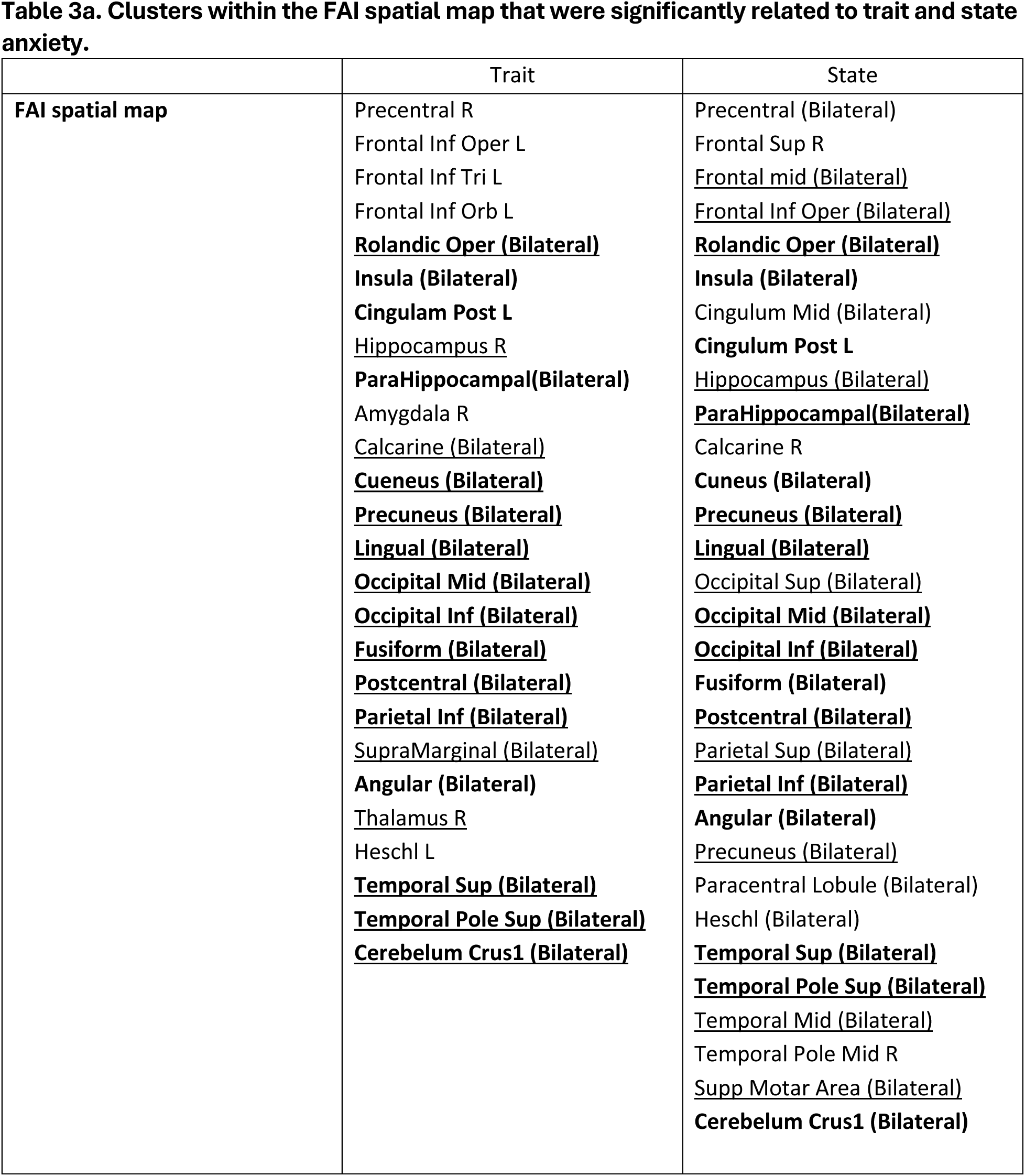
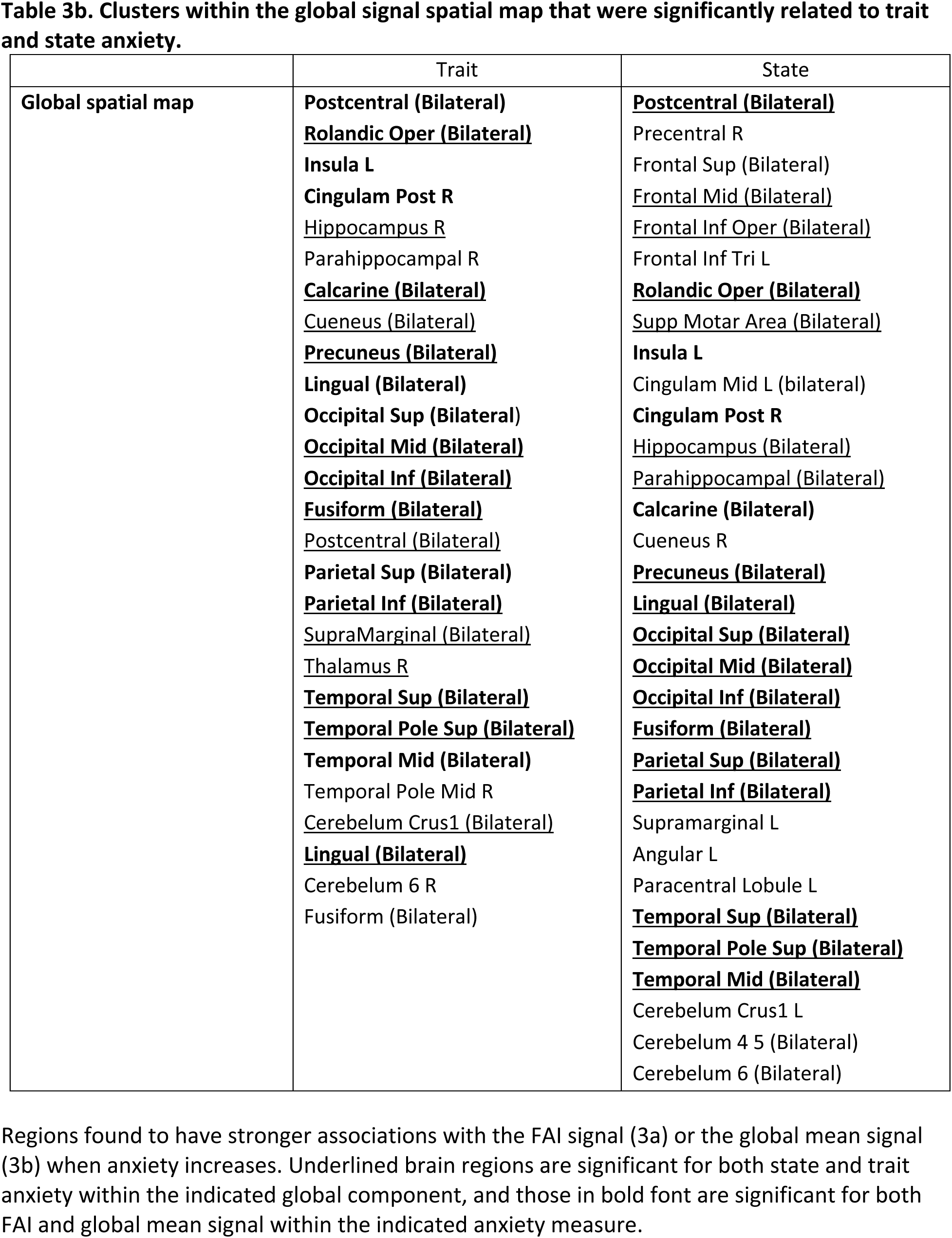
Clusters in which global components of the fMRI signal were associated with anxiety.

Regressing out the global mean signal weakened the relationship between VDMN-SAL and trait anxiety. In the subset of n=240 subjects with clean HR, the connectivity between DDMN-RCEN was related to trait anxiety at our uncorrected statistical threshold when none of the global components were regressed, and when HR or the FAI were regressed. However, regressing out the global mean signal removed this effect. These results did not change when age was not used as a covariate.

## Discussion

In a large community sample of adults, we demonstrate that global fMRI components linked with autonomic physiology and cortical arousal were significantly related to both state and trait anxiety measures. Additionally, regressing out the global mean signal and cortical arousal fluctuations altered the interaction between functional network connectivity and anxiety. Together, these findings suggest that global fMRI effects may be a source of information – and a novel avenue for identifying biomarkers of anxiety – as opposed to a confound^27^, and that careful consideration should be taken when determining whether to remove them in preprocessing.

We first tested if the brain-wide spatial distribution of the global mean signal, an fMRI cortical arousal index (FAI), and heart rate variation would relate to inter-individual differences in state and trait anxiety. We observed that anxiety measures were linked with clusters within maps associated to the global mean signal and FAI, but not heart rate. The superior and middle temporal pole, and bilateral precentral regions, were significant in all four comparisons relating the global mean signal or FAI with state or trait anxiety (**Tables 3a and 3b**). In other words, higher levels of anxiety corresponded to stronger associations between the global mean signal (or FAI).

In relating network signals with global component time-courses and anxiety, salience network emerged as exhibiting the most prominent relationships. First, the time-series of the salience network had the closest relationship with both the FAI and the global mean signal. These findings support the possibility that arousal may be crucial in the activation of the salience network at rest and potentially during cognitive tasks^43–45^. Second, the salience network’s interactions with the other networks of interest were most consistently related to anxiety measures at the selected threshold. The functional connectivity between the salience network and ventral default mode network decreased as both trait and state anxiety increased (p<0.05 uncorrected).

After regressing FAI from the network time-series, RCEN-SAL connectivity became more weakly related to state anxiety and VDMN-SAL connectivity became more weakly related to trait anxiety. Regressing out the global mean signal also reduced the relationship between VDMN-SAL connectivity and trait anxiety. These findings indicate that the global mean signal and the FAI may be capturing a component of the fMRI signal that relates certain network interactions to anxiety. However, unlike the relationships between global fMRI components and anxiety (**Figure 1**), none of the relationships between network connectivity and anxiety survived correction for multiple comparisons. Thus, caution should be taken when interpreting the present connectivity results and the impact of regressing out global components.

This study has several limitations. First, the community sample was not clinical, and the STAI measures were taken one day after the fMRI scan. These factors could potentially explain our observation that the relationship between network connectivity and anxiety levels was weaker and more variable than in certain patient studies. Second, the FAI reflects an estimated contribution of cortical arousal to fMRI signals. Simultaneous EEG or pupillometry and fMRI may more directly determine the extent to which cortical arousal effects relate to anxiety and/or contribute to network connectivity in anxiety disorders. In addition, fMRI measures and anxiety scores were examined across individuals, and future studies may also examine within-subject relationships. Finally, our findings may be limited to the resting-state experimental environment. The interplay between HRV and brain networks may be more dynamic during real-world conditions^46^, which may also explain why heart rate did not relate to anxiety in the present study.

To conclude, we find that global effects in fMRI, including those linked with cortical arousal, are closely tied with anxiety measures. These findings support the emerging viewpoint that global components – which are typically regarded as confounds in fMRI studies – may hold valuable information related to anxiety and perhaps to a broader range of clinical questions. We encourage future investigation into autonomic physiological or arousal measures in fMRI studies as potential predictive markers or within the context of mechanistic models.

## Data Availability

All data produced are available online at https://fcon_1000.projects.nitrc.org/indi/enhanced/access.html

https://fcon_1000.projects.nitrc.org/indi/enhanced/access.html

## Data Sharing Statement

This study used data from the Enhanced Nathan Kline Institute - Rockland Sample. The neuroimaging data from this sample is publicly available, and phenotypic data are available with a Data Use Agreement from the NKI. Code for reproducing this study will be shared no later than the date of manuscript acceptance.

## Acknowledgments

This work was supported by NIH grants T32 MH064913 (KRO) and T32AG058524/F99AG079810 (SEG). The study sponsors had no role in this work. This project drew upon publicly available data from the Enhanced Nathan Kline Rockland Institute Sample. The authors would also like to thank Justin Shao, for being a mentee who was eager to learn about different aspects of the project in its beginning stages. Additionally, we would like to acknowledge Dr. Allison Leich Hilbun for her consultation on the statistical sections of our work. Parts of this work were presented in abstract form at the 2024 meetings of the Organization for Human Brain Mapping and the Society for Biological Psychiatry. Author contributions: KRO - conceptualization, data curation, formal analysis, investigation, methodology, project administration, visualization, Writing—original draft, Writing—review & editing; CM - conceptualization, formal analysis, investigation, methodology, project administration; TL-Formal analysis, Visualization, Writing– original draft, and Writing–review & editing; SG - Software, Methodology. SW - Data curation, Methodology; RS - Data curation, Methodology; RY - Software, Data curation, Methodology; YL - Data curation, Methodology; JMH - Data curation, Methodology; RG - Software, RGB - Software, LQU - Writing-review & editing, MW - Writing-review & editing, JH - Resources, Investigation, Writing-review & editing; CC - Conceptualization, Methodology, Project Administration, Resources, Visualization, Writing–Original draft, and Writing–review & editing.

## Disclosures

K.R.O- no disclosures

C.G.M. - no disclosures

T.L. - no disclosures

S.E.G. - no disclosures

S.W - no disclosures

R.S. - no disclosures

R.Y. - no disclosures

Y.L. - no disclosures

J.M.H - no disclosures

R.G. - no disclosures

J.H. - no disclosures

R.G.B. - no disclosures

L.Q.U. - no disclosures

M.W.- is a member of the advisory boards and gave presentations for the following companies: HMNC, Janssen Pharmaceutical Research, Novartis, Boehringer Ingelheim, Germany; Bayer AG, Germany; and Biologische Heilmittel Heel GmbH, Germany. MW has further conducted studies with institutional research support from HEEL and from Janssen Pharmaceutical Research for a clinical trial (IIT) on ketamine in patients with major depression unrelated to this investigation.

C.C. - no disclosures

## Supplementary Material

**Supplementary Table 1.**
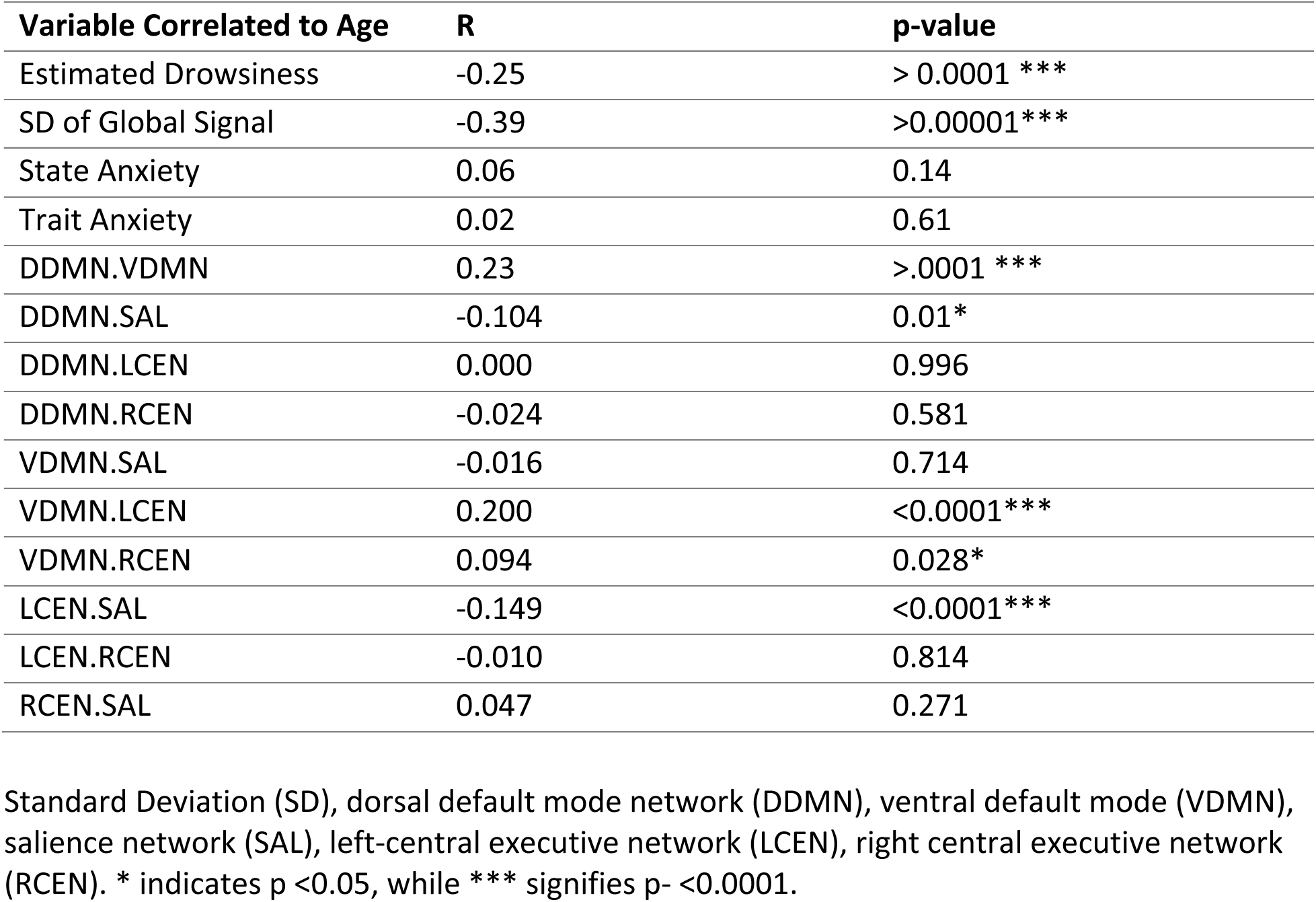
Correlation between age and several variables of interest.

**Supplementary Figure 1.**
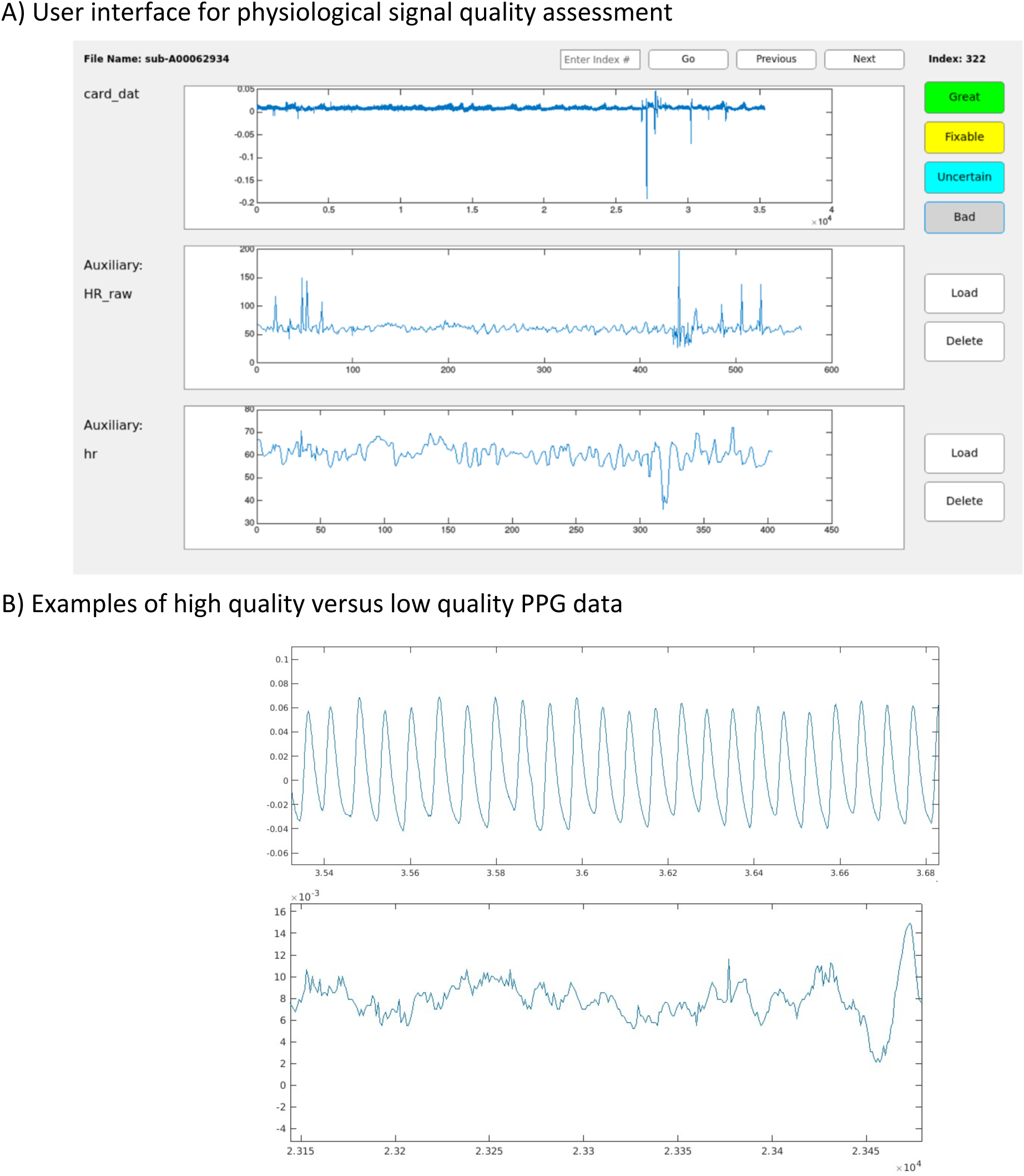
Manual inspection of physiological data quality. Top: The user interface for manual inspection of PPG data. We inspected three signals: raw PPG data (card_dat), an instantaneous heart rate measure (inter-beat-interval series, converted to HR by taking the inverse and multiplying by 60; HR_raw) and lastly, a heart rate fMRI regressor sampled at each TR (hr). B) An example of high vs. low quality PPG data.

**Supplementary Figure 2.**
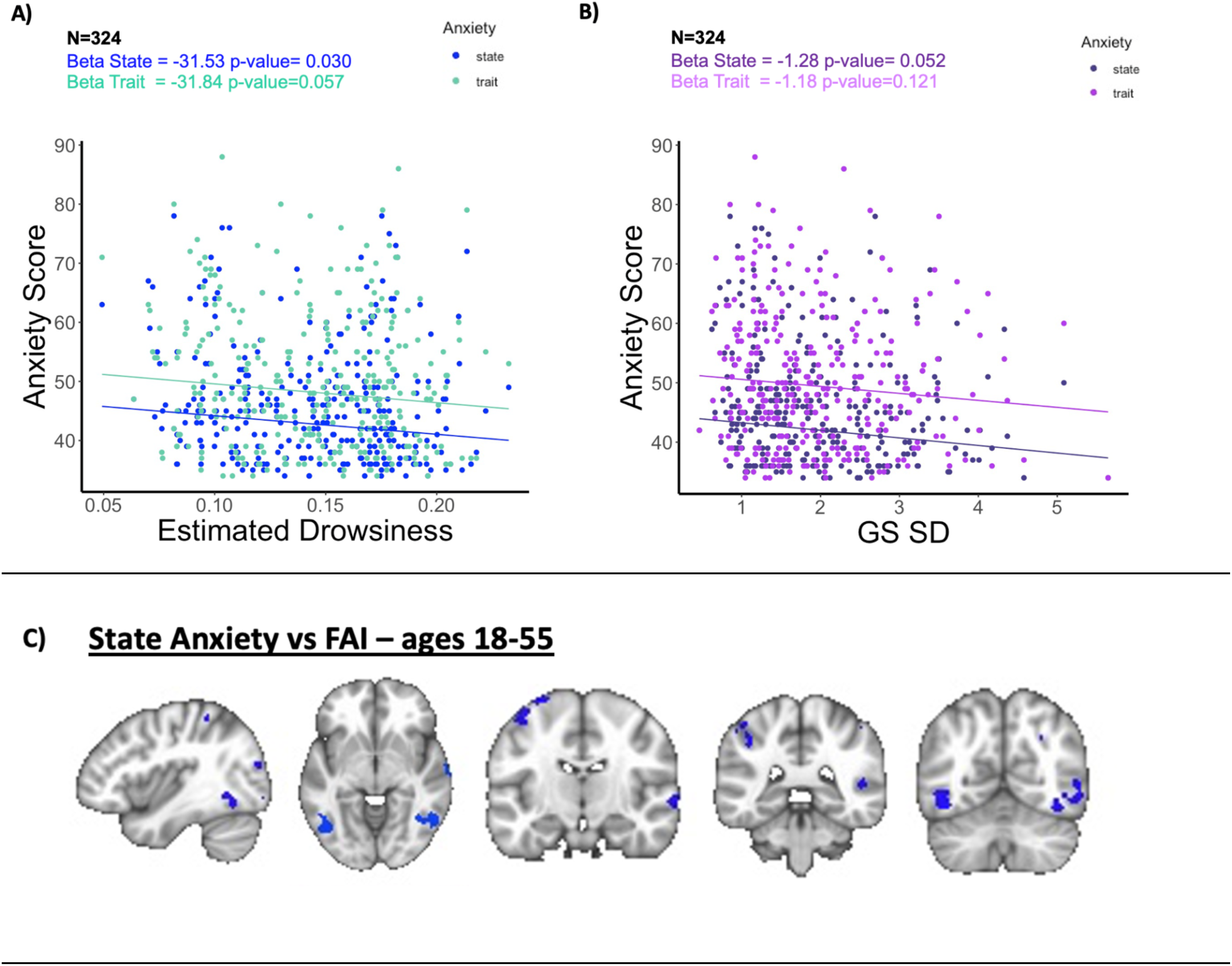
Relating spatial and dynamic aspects of global brain arousal effects to state and trait anxiety, in the set of participants aged 18-55. (A) Relationship between anxiety scores and the standard deviation of the global mean signal during the fMRI scans, covarying for age, sex, and ethnicity, in subjects aged 18-55. (B) Relationship between anxiety scores and the standard deviation of the arousal signal, covarying for age, sex, and ethnicity, in subjects aged 18-55. (C) Brain regions where subject-specific spatial maps associated with the FAI were significantly related to state anxiety, covarying for age, sex, and ethnicity (p<0.05, corrected for multiple comparisons using Threshold-Free Cluster Enhancement).

**Supplementary Figure 3.**
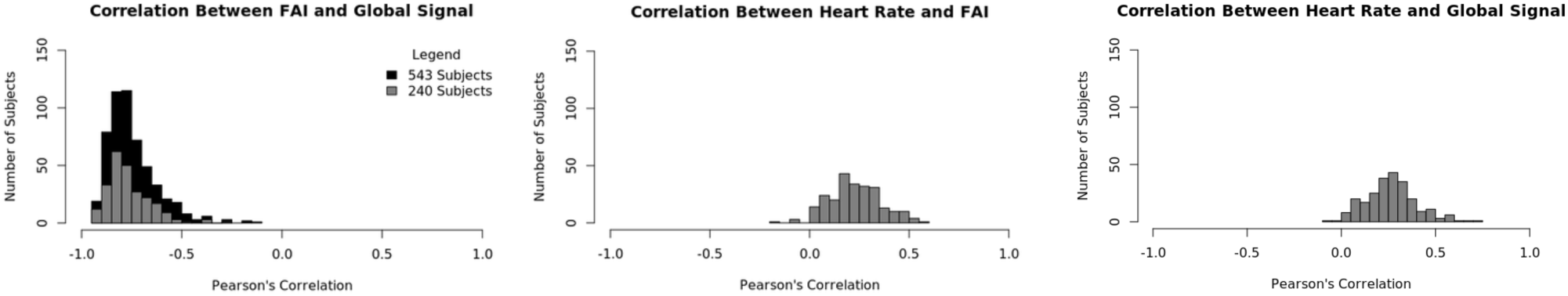
Correlations between the time courses of the indicated global components, for the sets of 543 subjects (black) and 240 subjects (gray)

**Supplementary Table 2.**
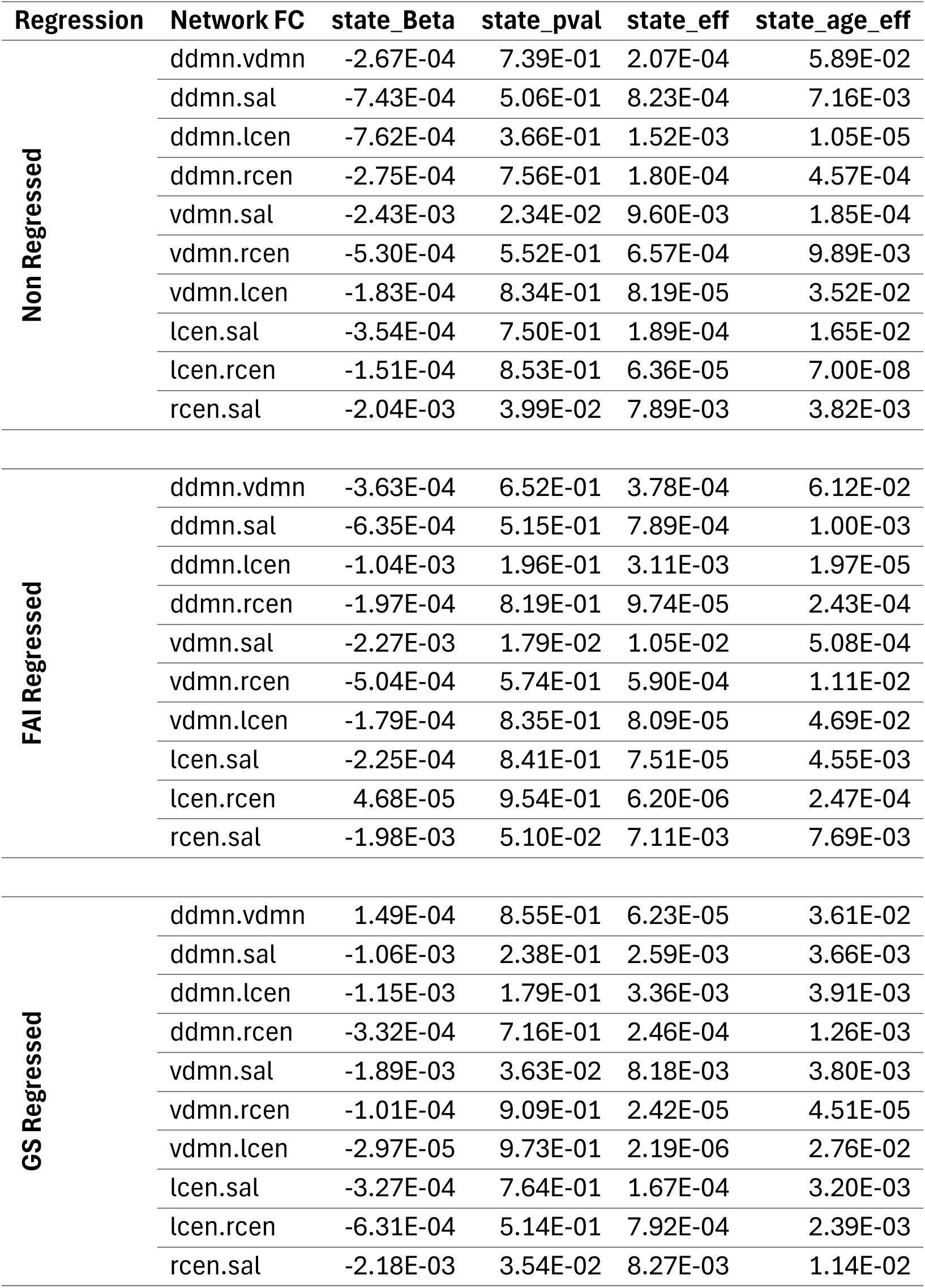
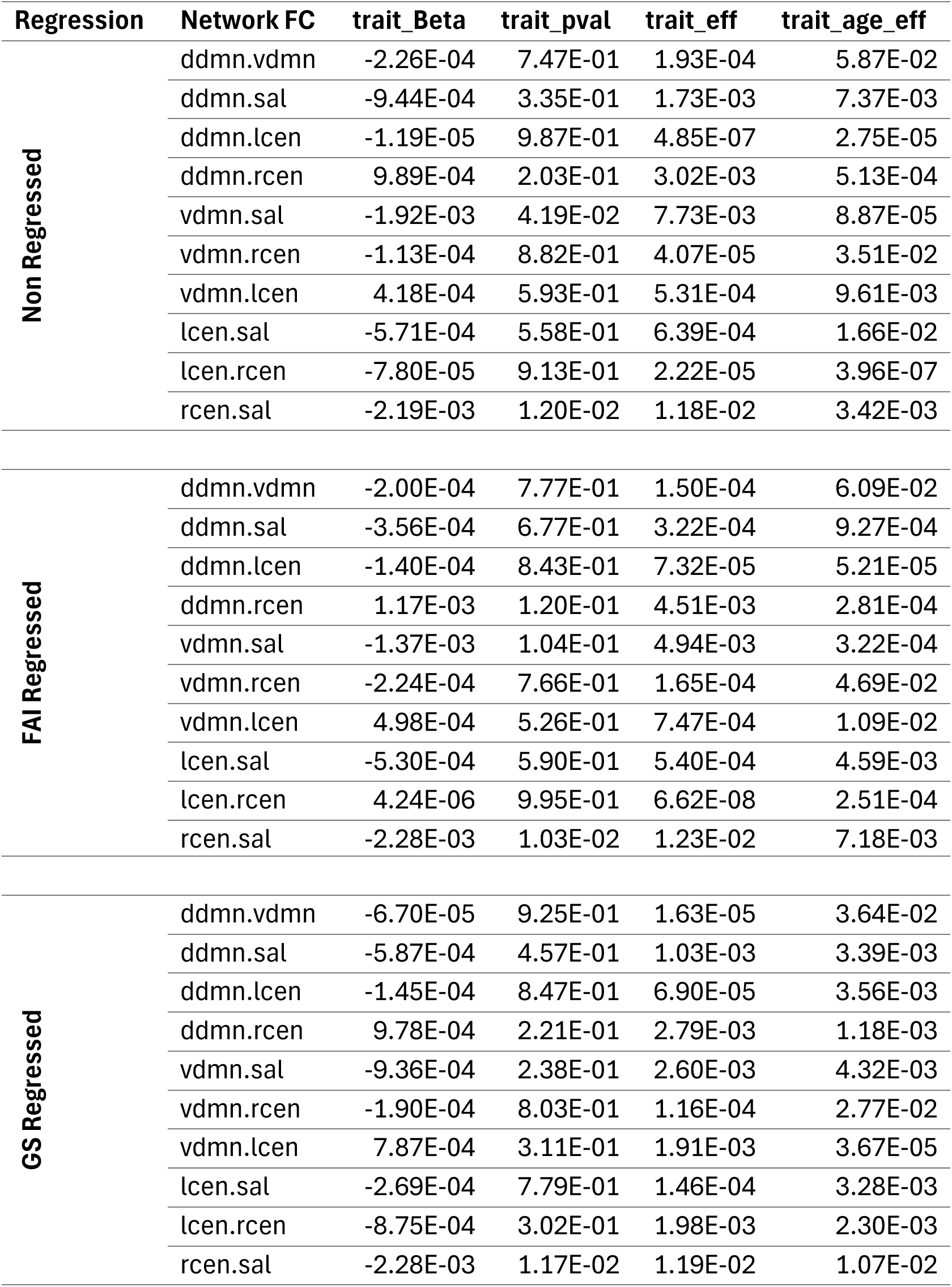
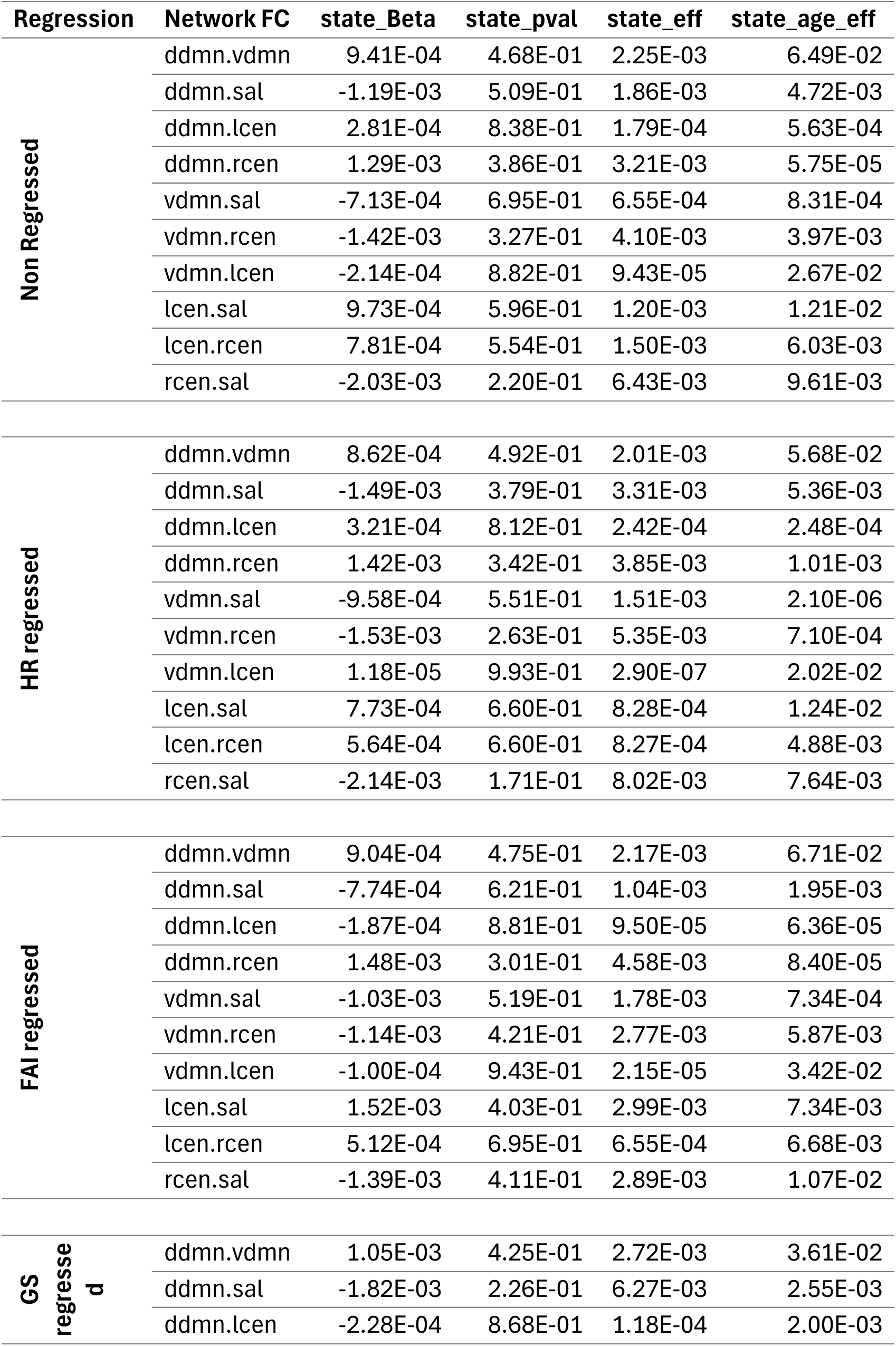
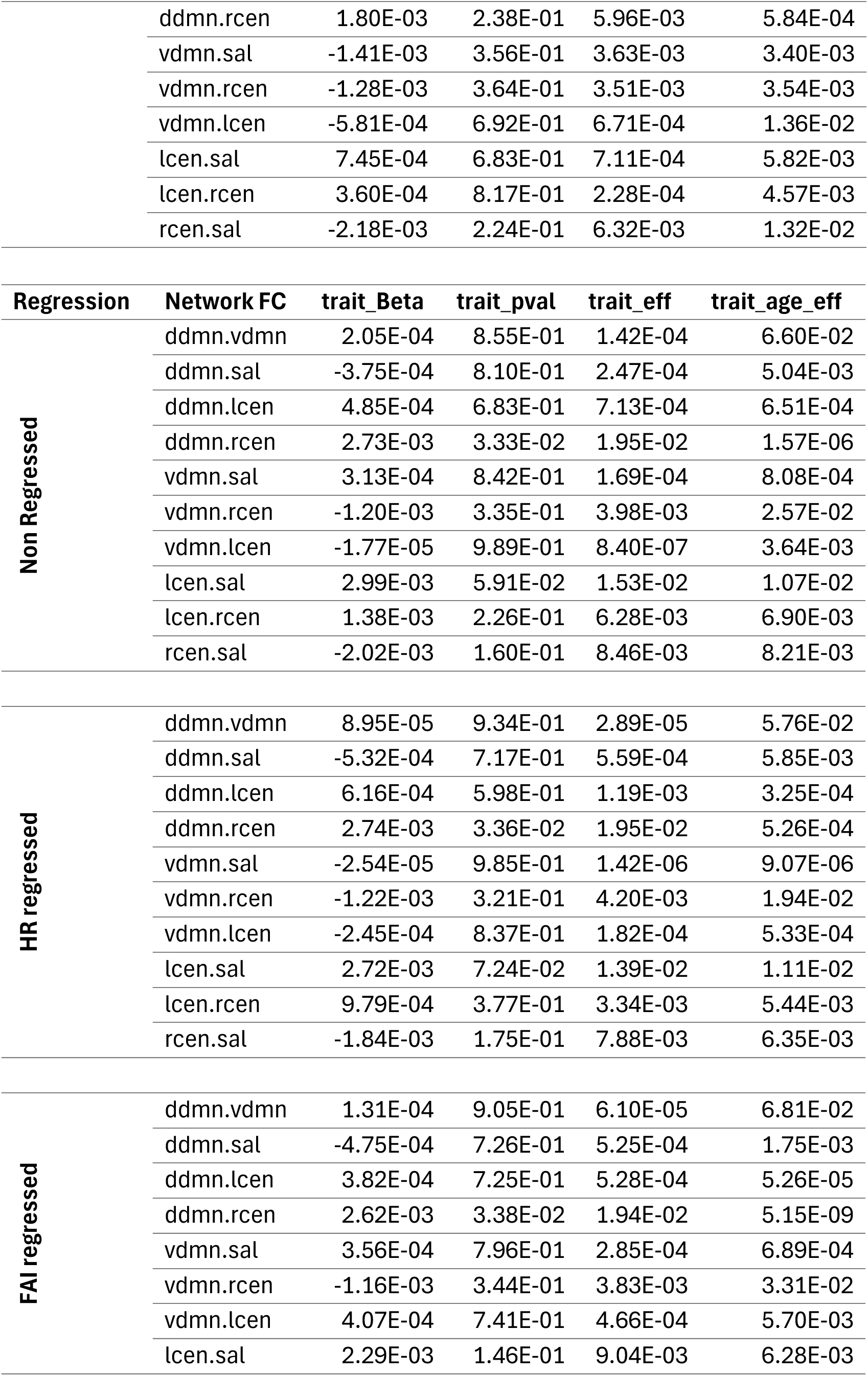
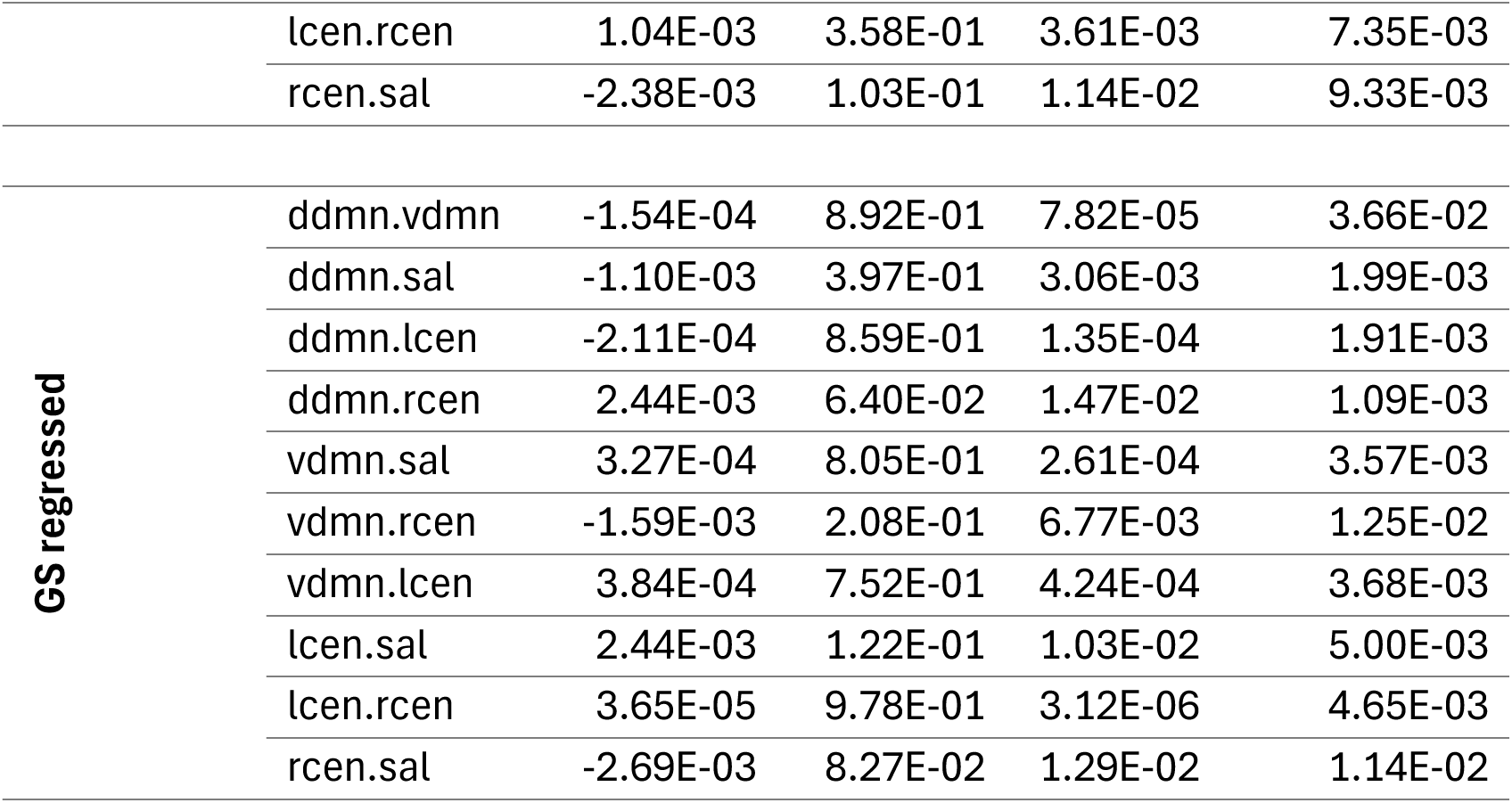
**a** Impact of regressing out global components on the relationship between network FC and state or trait anxiety (543 subjects). **b.** Impact of regressing out global components on the relationship between network FC and state or trait anxiety (240 subjects).

## Notes

### Funding Statement

This study was funded by NIH grants T32 MH064913 (KRO) and T32AG058524/F99AG079810

### Author Declarations

The study used openly available data set from the Nathan Klein Institute Rockland sample. The fMRI and heart rate measures were fully open source and available publicly. We did apply to receive participants state and trait anxiety scores. Ethics committee/IRB of Nathan Klein Institute gave ethical approval for us to download and use the state and trait data for our investigation.

